# Inactivated trivalent influenza vaccine is associated with lower mortality among Covid-19 patients in Brazil

**DOI:** 10.1101/2020.06.29.20142505

**Authors:** Günther Fink, Nina Orlova-Fink, Tobias Schindler, Sandra Grisi, Ana Paula Ferrer, Claudia Daubenberger, Alexandra Brentani

## Abstract

We analyzed data from 92,664 clinically and molecularly confirmed Covid-19 cases in Brazil to understand the potential associations between influenza vaccination and Covid-19 outcomes. Controlling for health facility of treatment, comorbidities as well as an extensive range of sociodemographic factors, we show that patients who received a recent influenza vaccine experienced on average 8% lower odds of needing intensive care treatment (95% CIs [0.86, 0.99]), 18% lower odds of requiring invasive respiratory support (0.74, 0.88) and 17% lower odds of death (0.75, 0.89). Large scale promotion of influenza vaccines seems advisable, especially in populations at high risk of severe SARS-CoV-2 infection.

**One Sentence Summary:** Covid-19 patients with recent influenza vaccination experience better health outcomes than non-vaccinated patients in Brazil.

## Main Text

As of June 12^th^, 2020, coronavirus disease 2019 (Covid-19) has affected 7.7 million individuals globally and caused an estimated 424,000 deaths. While the number of new Covid-19 cases has stabilized in most countries in the Northern hemisphere, the pandemic is expanding rapidly in the global South (*1*). Even though SARS-CoV-2 infections can be asymptomatic or result in mild disease in many cases, elderly individuals as well as individuals with pre-existing health conditions like obesity, cardiovascular diseases and diabetes mellitus are likely to develop serious and often life-threatening illness (*2, 3*). Currently, the mainstay of combating this novel virus is based on drastic public health measures affecting free movement of people and goods (*1*). This bold and unprecedented approach has resulted in large scale lockdowns of entire countries, potentially resulting in the largest global economic downturn since World War II (*4*). In the absence of an effective drug treatment (*5*) and without a validated vaccine against SARS-CoV-2 (*6*) public health interventions with beneficial impact on Covid-19 outcomes, particularly for the most vulnerable populations, are urgently needed.

Many countries south of the equator are now entering the cold season of the year, which will likely result in a substantial increase in patient volume due to influenza. Seasonal influenza outbreaks occur in regular intervals in most non-tropical countries, and are estimated to cause 650,000 deaths each year (*7*). Despite major public health efforts, influenza vaccines remain under-used in most countries, mostly due to skepticism regarding their efficacy and concerns surrounding their safety (*8, 9*). Several social media rumors linking influenza vaccine to adverse Covid-19 outcomes in recent weeks (*10*) like have further undermined the willingness of the general population to undergo influenza vaccination.

In this paper, we use detailed medical records from over 90,000 clinical Covid-19 patients in Brazil to compare Covid-19 outcomes of patients with and without recent influenza vaccination. As of June 12, Brazil is the country with the second highest number of Covid-19 cases, as well as the country with the largest number of new cases documented each day.

As described in further detail in **Supplemental Materials and Methods**, all Covid-19 cases identified at Brazilian health centers have to be reported mandatorily into a central database system. As of June 9^th^ 2020, 92664 clinically confirmed Covid-19 patients were registered in this system. A positive laboratory test for SARS-CoV-2 was documented in the medical records for 84% of the clinically diagnosed Covid-19 patients. 57% of patients were male, and the median age of patients was 59 years (**Table 1**). The most represented age group among Covid-19 patients were individuals between 60 and 69 years of age. 37.1% of patients required intensive care, 22.5% were given invasive respiratory support, and 47.1% of patients died. Covid-19 fatality rates increased from 15% among children under the age of 10 to 83% among individuals above 90 (**Supplementary Materials Figure S1**). 66.4% of Covid-19 patients had a preexisting cardiovascular condition and 54.9% has previously been diagnosed with diabetes mellitus. The prevalence of obesity, neurological disorders and renal disease were 10.9%, 11.0% and 11.9%, respectively (**Supplementary Materials Figure S2**).

**Table 1:** Severity and mortality outcomes of Covid-19 patients by gender, age, race andeducational attainment.

|  | N | % | Proportion with influenza vaccine | Proportion in intensive care | Proportion with invasive respiratory support | Proportion deceased |
| --- | --- | --- | --- | --- | --- | --- |
| Total | 92'664 | 100% | 0.311 | 0.371 | 0.225 | 0.471 |
| Male | 53'005 | 57.2% | 0.284 | 0.383 | 0.236 | 0.483 |
| Female | 39'659 | 42.8% | 0.348 | 0.356 | 0.211 | 0.456 |
| Age 0-9 | 1'175 | 1.3% | 0.181 | 0.337 | 0.156 | 0.161 |
| Age 10-19 | 740 | 0.8% | 0.141 | 0.293 | 0.142 | 0.190 |
| Age 20-29 | 3'846 | 4.2% | 0.222 | 0.247 | 0.111 | 0.139 |
| Age 30-39 | 10'269 | 11.1% | 0.233 | 0.280 | 0.116 | 0.169 |
| Age 40-49 | 14'390 | 15.5% | 0.216 | 0.307 | 0.154 | 0.253 |
| Age 50-59 | 16'965 | 18.3% | 0.227 | 0.358 | 0.213 | 0.375 |
| Age 60-69 | 17'746 | 19.2% | 0.426 | 0.408 | 0.283 | 0.557 |
| Age 70-79 | 14'826 | 16.0% | 0.438 | 0.441 | 0.313 | 0.678 |
| Age 80-89 | 9'861 | 10.6% | 0.439 | 0.450 | 0.295 | 0.769 |
| Age 90+ | 2'846 | 3.1% | 0.410 | 0.424 | 0.214 | 0.846 |
| White | 26'526 | 28.6% | 0.359 | 0.378 | 0.206 | 0.400 |
| Mixed | 27'417 | 29.6% | 0.269 | 0.310 | 0.228 | 0.548 |
| Black | 4'259 | 4.6% | 0.346 | 0.322 | 0.216 | 0.491 |
| Other/NA | 34'462 | 37.2% | 0.292 | 0.422 | 0.241 | 0.465 |
| No education | 1'980 | 2.1% | 0.398 | 0.241 | 0.202 | 0.716 |
| Primary | 7'058 | 7.6% | 0.381 | 0.280 | 0.206 | 0.578 |
| Lower secondary | 5'190 | 5.6% | 0.286 | 0.309 | 0.207 | 0.473 |
| Upper secondary | 10'414 | 11.2% | 0.292 | 0.305 | 0.166 | 0.341 |
| Higher education | 5'811 | 6.3% | 0.416 | 0.372 | 0.146 | 0.236 |
| Still in school | 770 | 0.8% | 0.176 | 0.353 | 0.181 | 0.174 |
| Not available | 61'441 | 66.3% | 0.279 | 0.404 | 0.249 | 0.498 |

31.1% of Covid-19 patients had received an influenza vaccine during the last campaign, with highest rates among individuals aged 60 and older, as well as among people with higher education. The Brazilian Ministry of Health (MoH) has been conducting annual vaccination campaigns achieving relatively high population coverage since 1999 (*11, 12*).

Seasonal influenza in Brazil peaks in weeks 18-19 (April-May) in northern states and in weeks 25-27 (June-July) in southern states (*13*). The 2020 annual influenza vaccination campaign was launched on March 23^rd^, one month earlier than originally planned to ensure vaccine delivery to the public prior to the incoming wave of SARS-CoV-2 infections, with the ambition to reach a total of 67.6 million people nationwide (*14*). The national campaign targets senior citizens (ages 60 and older) and health workers in phase 1; chronic patients with comorbidities and chronic disease conditions, teachers, security and rescue forces in phase 2; and children and other high risk populations in phase 3. Based on the recommendation from the WHO, a trivalent (type A/Brisbane/02/2018 - IVR-190 (H1N1), type A/South Australia/34/2019 - IVR-197 (H3N2) and type B/Washington /02/2019), non-adjuvant influenza vaccine produced in Brazil by the Instituto Butantan is currently used (*15*). **Supplementary materials Figure S3** illustrates vaccination coverage by age – rates were substantially higher among children under age 6 as well as adults aged 60 and above, but were below 50% in all age groups in our population analyzed here.

**Figure 1** shows mortality patterns by age and vaccination status. Covid-19 related mortality ranged from 14% among children under the age of 10 to 84% among individuals aged 90 or older in the non-vaccinated group. Mortality was consistently lower among influenza vaccinated patients across all age groups, with absolute mortality differences ranging from a risk difference of 17% pts in the 10-19 age group to a risk difference of 3% pts in the 90+ age group. This difference was statistically significant (p-value < 0.05) for all age groups over 30.

**Figure 1:**
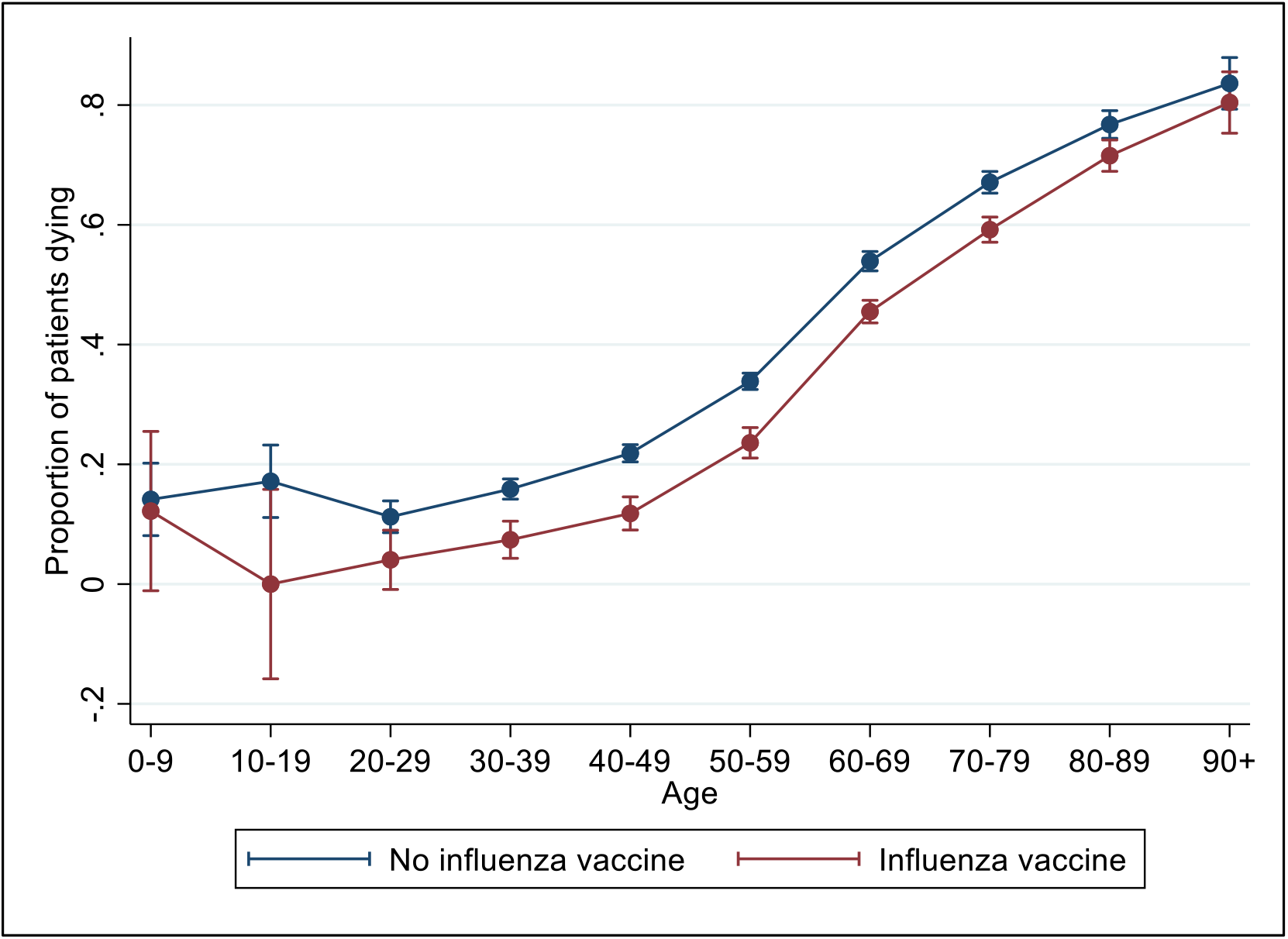
Covid-19 mortality by age and vaccination status. shows the proportion of Covid-19 patients dying by age group and influenza vaccination status. Estimates represent un-adjusted linear differences in age-group specific mortality with 95% confidence intervals.

To estimate the associations between influenza vaccination status and Covid-19 mortality, we used multivariable logistic regression models, conditioning on an increasing number of potential confounding factors. When we conditioned the model on age only, we find that influenza vaccination is associated with a 35% reduction in the odds of death among Covid-19 patients (column 1, **Table 2**). Given vaccination rates are likely higher in areas with more effective health systems, we further restrict our analysis to within-facility comparisons (columns 2-5, **Table 2)**. When we exclusively compare outcomes among patients getting care at the same facility, the protective association attenuates to 18% (p-value < 0.01). Given that even within facilities vaccinated patients may on average have better initial health, we also control for a full set of preexisting co-morbidities in a separate model – the estimated positive association remains nearly identical (column 3, **Table 2**). To address potential confounding concerns regarding health knowledge or general health behaviors (not manifested in the observed comorbidities), we also include socioeconomic controls such as gender, race and educational attainment in our full empirical model– once again, the estimated positive association changes only marginally (column 4, **Table 2**). The same associations are also observed when we restrict the cohort to patients with a positive SARS-CoV-2 specific RT-qPCR result documented in the medical records (i.e. when we exclude the 16% of the Covid-19 patient where a positive laboratory SARS-CoV-2 test result was not documented in the records).

**Table 2:**
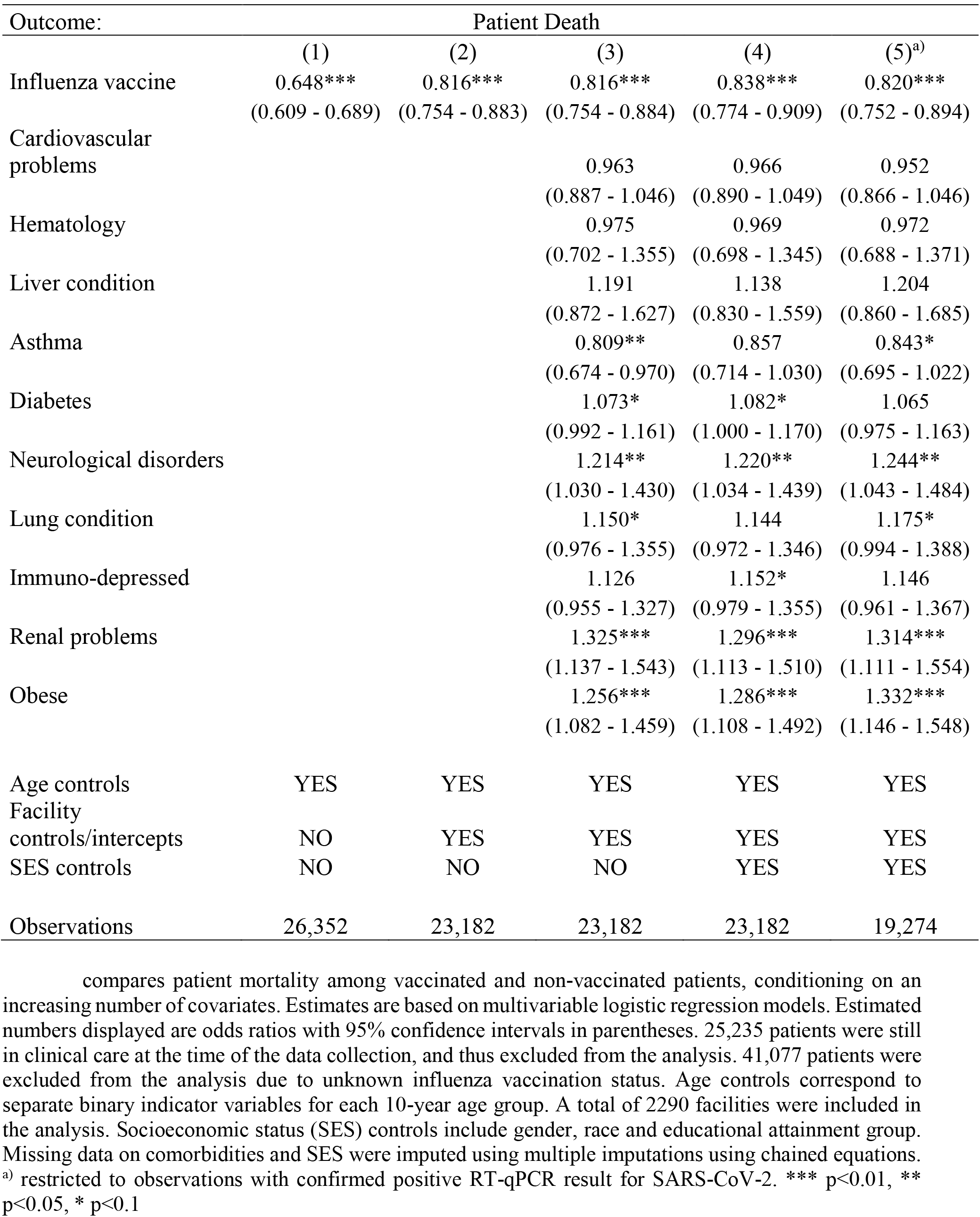
Estimated associations between influenza vaccination and Covid-19 mortality.

As expected, known risk factors for Covid-19 related mortality like obesity, pre-existing lung conditions, renal problems and neurological disorders were associated with higher mortality, with estimated odds ratios between 1.17 (lung conditions) and 1.33 (obesity). Patients suffering from asthma had marginally lower mortality odds (aOR 0.84, [0.695 - 1.022]) (**Table 2**).

**Table 3** shows estimated associations between vaccination status and clinical care received. As before, we restrict our analysis to within-facility comparisons and condition on the full set of socioeconomic and comorbidity controls. On average, influenza vaccination was associated with an 8% reduction in the odds of receiving intensive care, and a 19% reduction in the odds of receiving respiratory support. **Supplementary Materials Table 2** shows further details for all covariates included in this analysis.

**Table 3:** Estimated associations between vaccination status and Covid-19 severity.

| Outcome: | Intensive Care |  | Respiratory Support |  |
| --- | --- | --- | --- | --- |
|  | (1) | (2) | (3) | (4) |
| Influenza vaccine | 0.914**<br>(0.852 - 0.980) | 0.924**<br>(0.861 - 0.992) | 0.800***<br>(0.736 - 0.869) | 0.805***<br>(0.741 - 0.875) |
| Age controls | YES | YES | YES | YES |
| Facility controls/intercepts | YES | YES | YES | YES |
| SES & comorbidity controls | NO | YES | NO | YES |
| Observations | 26,260 | 26,260 | 25,959 | 25,959 |

Figure 2 shows estimated associations between vaccination status and Covid-19 mortality, separated by the timing of influenza vaccine administration. As shown in **Supplemental Materials Figure S4**, most of the vaccines (65.7%) were administered as part of the 2020 campaign, with 776 vaccinations (6.8%) administered after the self-reported on-set of Covid-19 symptoms. When we jointly analyze all age groups (Panel A, Figure 2), we find protective effects for individuals whose last vaccination was given in March 2020 or later, but not for individuals last vaccinated earlier. On average, we find that vaccines obtained as part of the 2020 campaigns were associated with 20% lower odds of mortality if the vaccine was given prior to the onset, and with 27% lower odds of death if the vaccine was given after onset of clinical Covid-19 symptoms – these two estimates are however not statistically different from each other.

**Figure 2:**
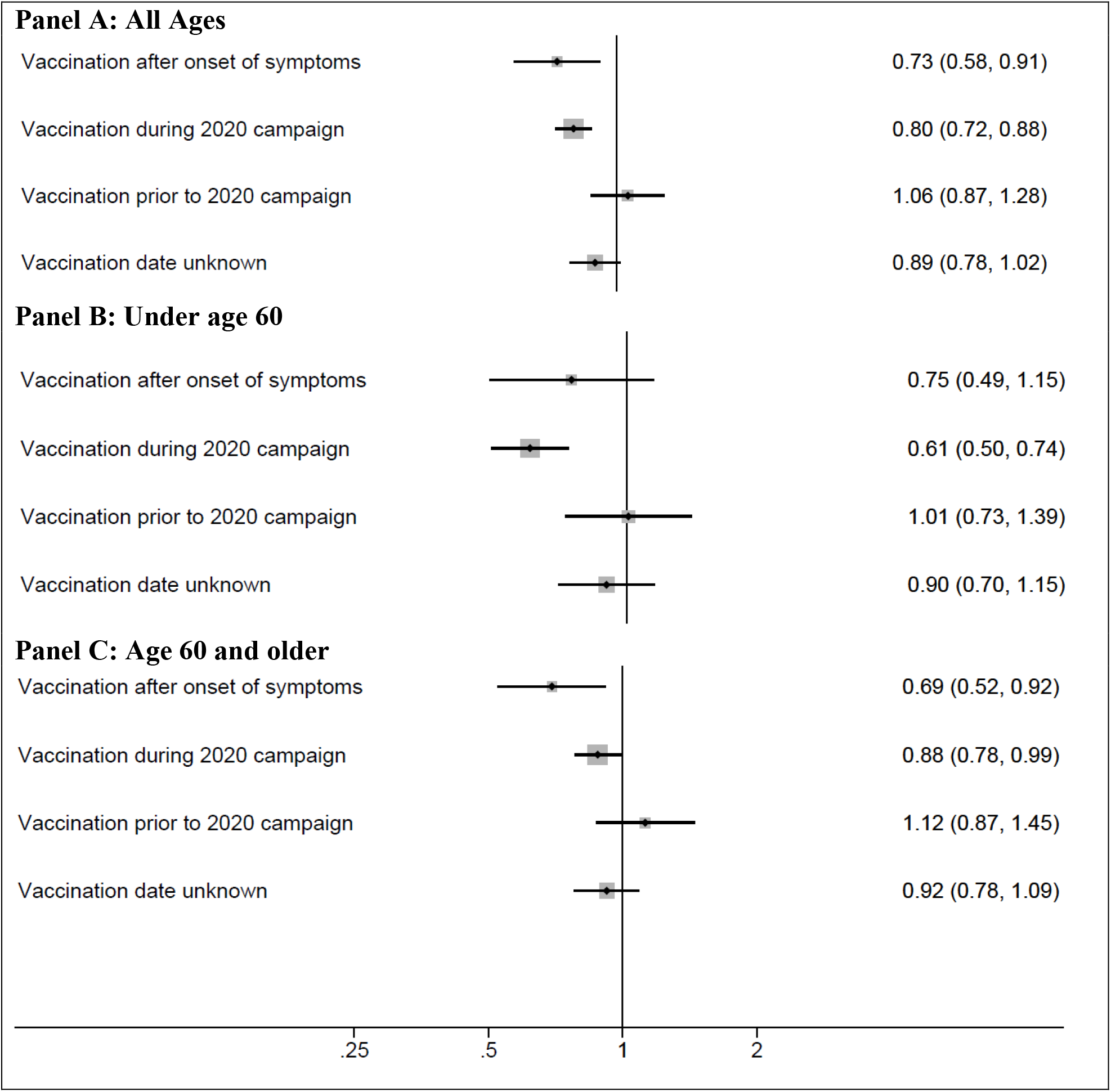
Estimated association between influenza vaccination status and Covid-19 mortality dependent on timing of vaccine administration and stratified by patient age. compares Covid-19 mortality outcomes of patients vaccinated against influenza at different points in time to Covid-19 mortality outcomes of non-vaccinated patients. Estimates represent odds ratios based on multivariable logistic regression models with full set of covariates (95% confidence intervals displayed as lines around the point estimate). The size of the grey squares is proportional to the sample size in each group. Panel A includes all patients; panels B and C include patients under and over the age of 60, respectively.

When we stratify our analysis by age (Panel B and C, Figure 2), we find larger protective effects for patients under the age of 60 than for older patients when the vaccine was given prior to onset of symptoms. For vaccines given after onset of symptoms, effect sizes in both age groups (under and above 60) seem very similar. No protective effects were found for either age group in cases when influenza vaccine was given prior to the current campaign, or when the date of last vaccination was unknown. The full regression results underlying these figures are provided in **Supplementary Materials Table 2**.

## Discussion

The results presented in this study strongly suggest that concerns regarding potential negative side effects of influenza vaccination in case of ongoing spread of SARS-CoV-2 infections are not warranted. We show that on average Covid-19 patients who received the inactivated trivalent influenza vaccine in 2020 – even if administered after the onset of SARS-CoV-2 infection-related symptoms – had significantly higher chances of surviving and less need for intensive hospital care than patients without recent influenza vaccination. Patients obtaining an influenza vaccine differ potentially from non-vaccinated patients in a range of possible factors including genetics, health status, health pursuing behavior or other unknown biological or environmental factors that could at least partially explain the observed differences in clinical presentation and survival. We addressed these concerns in multiple ways: (i) we restricted our comparisons to patients using the same health facility, which eliminates differences in health care access and quality of care; (ii) we controlled for race to address immunogenetic differences of populations; and (iii) we controlled for age, gender and educational attainment to account for general differences in living conditions and health behaviors. Lastly, and most importantly, we controlled for an extensive set of pre-existing comorbidities documented in the medical records, which allows us to rule out any confounding through pre-existing health conditions.

While we cannot rule out residual confounding through behavioral differences not manifested in acute or chronic health conditions, it seems somewhat unlikely that such confounders would fully explain the substantial protective effects observed. This naturally raises the question about the most likely mechanisms underlying a potential protective effects of influenza vaccines. The most immediate explanation that comes to mind is the prevention of potential influenza-Covid-19 coinfections (*16*). Although individual cases of such coinfections have been documented (*17-19*), larger studies have found this combination to be rather rare (*20-22*). Given that we found only 30 cases with such coinfections among the > 90,000 patients in our data set, we can mostly rule out coinfections as mechanism underlying the protective associations observed.

From an immunological perspective influenza vaccines are designed to induce adaptive, long-lasting pathogen-specific immunity through secretion of neutralizing antibodies and development of specific T-cell responses (*23*). Even though influenza and SARS-CoV-2 display only limited nucleotide sequence similarity overall, CD8+ T-cell epitopes with modest sequence resemblance seem plausible (*24*). However, given the extraordinary diversity of influenza viruses, the induction of cross-neutralizing antibodies and T-cells that directly target other RNA viruses like SARS-CoV-2 seems somewhat unlikely. If such a long-lasting immunity had been the main mechanism of cross-protection, we should have also observed similarly protective effects for Covid-19 patients vaccinated in prior years, which was not the case in our analysis.

In our view, the most plausible mechanism underlying the beneficial effects observed are changes in innate immunity. A growing body of evidence has documented that immunological memory is not an exclusive feature of the adaptive immune response, and that adaptive features like memory can also be found in innate immune cells as well as in tissue-resident stem cells (*25*). These innate immune mechanisms can be triggered both by natural infections and by vaccines (*26, 27*) and result in “off-target” protective effects against a range of pathogens not directly aimed at by the vaccine (*28*). In vitro re-stimulation of peripheral blood leucocytes after BCG or influenza vaccination with a variety of unrelated antigens has been shown to result in a broadly modified cytokine profile with enhanced TNF-a and IL-6 production (*29, 30*). Observational, epidemiological and clinical studies have also provided strong evidence that live vaccines against *Mycobacterium tuberculosis* (BCG), measles, smallpox and polio can yield substantial non-specific protective effects against other pathogens (*31*), and that these off-target effects account for a significant share of the overall mortality reductions achieved by these vaccines (*32*). Given the high similarities of SARS-CoV-2 and influenza viruses with respect to viral structure, transmission and pathogenic mechanisms, it seems plausible that both viruses are detected by similar or identical pattern recognition receptors. Their binding to viral RNA can then trigger suitable inflammatory and antiviral responses. Two recent studies suggests that the immunogenicity of influenza vaccines depends on the binding of single stranded RNA to toll like receptor 7 (*33, 34*), which results not only in increased neutralizing antibody titers and T-cell activation, but also in enhanced activation of natural killer (NK) cells and early respiratory IL-12p40 and IFN-I responses (*33, 35*). A recent study also suggests that influenza vaccines can prime myeloid cells for enhanced cytokine secretion for up to 30 days, with lasting functional change in the NK cell subsets (*36*). These trained, memory like NK cells could potentially be stimulated by other RNA viruses including SARS-CoV-2 as well. These proposed mechanisms appear consistent with the differential protective effects observed in our study for the older patient group. Influenza vaccines generally appear less effective in older than in younger individuals (*37*). It thus seems plausible that innate immune system adaptations after vaccination may be compromised by aging and age-associated immune dysfunctions (*38*).

A surprising finding in our analysis is that influenza vaccination conducted at the time of onset of Covid-19 clinical symptoms or shortly thereafter was still associated with improved health outcomes. It is possible that the innate immune response induced by such late vaccination results in (i) more rapid and efficient SARS-CoV-2 clearance, preventing progressive dissemination into lower areas of lung tissues and/or (ii) dampening of the uncontrolled, destructive pro-inflammatory host response seen in Covid-19 at later, often fatal disease stages. It will be of paramount importance to more closely study these potential off-target effects of influenza vaccination in future research to fully understand the cell subsets affected, as well as the duration, reversibility and consequences of those changes on local immune defense mechanisms in the upper and lower respiratory tract.

Here, we provide strong, and to our knowledge first evidence that people at risk of developing severe Covid-19 disease might benefit significantly from influenza vaccination. We believe these off-target protective effects of influenza vaccines are most likely driven by a rapidly induced, altered activation stage of the innate immunity. This activation can either down-modulate immune overdrive induced by SARS-CoV-2 or directly prevent viral dissemination to the lower respiratory tract (*39*). In the absence of a Covid-19 vaccine and without a well-established treatment to avert disease progression, induction of trained immunity exerting beneficial, off-target effects might be a fruitful avenue for improving Covid-19 outcomes. The ongoing clinical studies with BCG are a promising step in this direction (*40*).

Given the proven benefits of influenza vaccination in terms of reduced influenza incidence (*41*), the incentives for governments and health care providers to aggressively promote seasonal influenza vaccination during the ongoing Covid-19 pandemic seem stronger than ever. Even if only a limited reduction in Covid-19 fatalities could be directly achieved by influenza vaccination, the absolute numbers of saved lives might be significant. Reducing the overall burden of respiratory viral infections on health care facilities during the influenza season will without any doubt alleviate the work load on the already strained health care work forces and will allow health systems to free and preserve care capacity for patients in greatest need until an effective vaccine or drug against SARS-CoV-2 can be developed and widely distributed.

## Data Availability

All data used in this analysis is publicly available through the Brazilian Ministry of Health webpage at https://opendatasus.saude.gov.br/dataset/bd-srag-2020. Code to replicate the analysis is available on request from the first author.

## Acknowledgments

None.

## Author contributions

GF, AB, CD and NO conceptualized the paper. GF conducted the analysis and created a first draft. All coauthors provided input on the multiple draft versions and approved the final version of the manuscript.

## Data and materials availability

All data used in this analysis is publicly available through the Brazilian Ministry of Health’s webpage at https://opendatasus.saude.gov.br/dataset/bd-srag-2020. Code to replicate the analysis is available on request from the first author.

## Supplementary Materials

### Materials and Methods

#### Study design

The study was designed as a clinical cohort study following all clinical patients with confirmed Covid-19 diagnosis registered in Brazil between January 29, 2020 and June 8, 2020.

#### Setting

The study was conducted in Brazil. Brazil is the sixth-most populous country in the world, with an estimated population of 212 million in 2019. As of June 12, Brazil is the country with the second highest number of Covid-19 cases, as well as the country with the largest number of new cases documented each day (https://coronavirus.jhu.edu/map.html=

#### Participants

All individuals with a severe respiratory infection registered in the health system between January 1, 2020 and June 8, 2020 were analyzed. Patients whose final survival outcome was not documented as of June 8 were excluded from the mortality analysis.

#### Variables

Our main outcome of interest is progression to severe disease. Our primary outcome variable was patient survival; we also analyzed intensive care treatment as well as invasive respiratory support as (intermediate) secondary outcomes.

Our primary independent or exposure variable of interest was vaccination status. The national reporting system requires providers to complete a detailed patient report that is designed to capture basic demographics, symptoms and co-morbidities. Given Brazil’s major commitment to national vaccination campaigns, the form also contains a question on whether the patient obtained an influenza vaccine as part of the ongoing national vaccination campaign. If the patient confirms this, the vaccination date e is also entered in the system.

#### Data sources

All data used was collected within the Brazilian surveillance system for severe respiratory infections (“*Vigilância de Síndrome Respiratória Aguda Grave*” (SRAG)). Since 2009, Brazil operates a national disease surveillance network, which requires all health facilities and providers to report all infections disease cases into a central system using a standardized reporting protocol. The Covid-19 pandemic is considered a national public health emergency, which requires that all cases must be reported within 24 hours to the MoH. All major respiratory infections are then captured in a central system (SISRAG – Sistema de Informação de Síndrome Respiratória Aguda Grave). In compliance with Brazilian public law (Lei 12.527/2011, art. 7, § 3°), the MoH makes these surveillance data publicly available, after removing all identifiable information. Even though providers are formally required to complete forms for all patients, some fields (such as symptoms and co-morbidities) are not always filled.

#### Bias

The primary bias concern for our analysis is confounding: given that the national campaign targets children and older individuals, age is an obvious concern; this can easily be addressed by adjusting all estimates for age. It also seems possible that vaccines are more common among individuals that are health workers or work-related target population, more health-aware people, or have better health access. To address this, we control for educational attainment as well as facility of treatment in our adjusted models. Data on occupation was not available in the database.

#### Study size

The study included all cases of severe respiratory infection documented between January1 and June 8, 2020. The data set was downloaded on June 9, 2020.

#### Statistical methods

We start by presenting sociodemographic characteristics of patients, as well as the proportion of patients requiring intensive care and respiratory care by gender, age, education and race. We also show average survival probabilities for this group. In a second step, we plot average survival probabilities for vaccinated and non-vaccinated individuals by 10-year age group.

In a third step, we use multivariable regression models to estimate the associations between vaccination status and health outcomes. We first show basic correlations between influenza vaccination status and outcomes. To address the most obvious confounding concerns (age and health care access), we next show empirical models that control for age and use within-facility variation only by including facility-specific intercepts (fixed effects) in our empirical models. To further control for potential differences in disease severity, we show models that control for an extensive list of co-morbidities documented in the data. Last, we also show models that control for educational attainment and race to address concerns regarding selective vaccination uptake within facilities. All main figures are presented in the text – additional materials are provided below.

#### Missing Data

The final survival outcome (survived or died) was available for 67429 out of 92664 patients (72.8%). Vaccination status was available for 36650 out of 92664 cases (39.6%). About 40% of cases with undefined vaccination status was explained in reporting differences across facilities. The remaining variation in reporting reflects differences in individual provider efforts to enter complete patient data into the system. Given the focus of this paper, we focused on the subset of patients where vaccination data was available. To address missingness in the covariates included in the analysis (particularly for comorbidities and socioeconomic variables) multiple imputation using chained equations was applied. We used Stata’s MI package to run these imputations – the final results were based on 100 random imputations.

**Supplementary Materials Figure S1:**
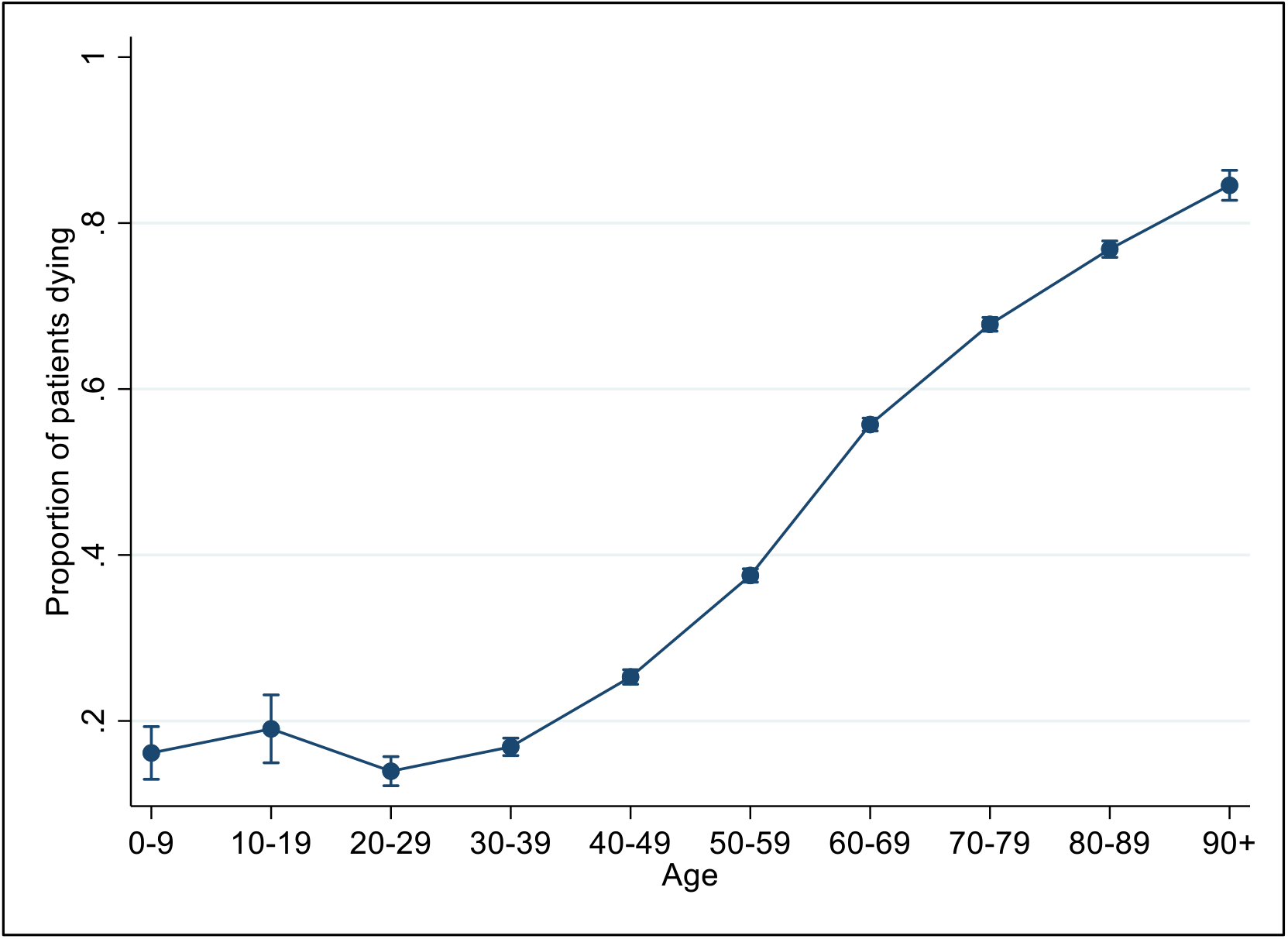
Covid-19 mortality by age group. shows age specific mortality rates by 10-year age group. Mortality on the y-axis is expressed as proportion of Covid-19 patients in each age group dying. Analysis was restricted to patients where the final outcome (survived or died) was known at the time of data extraction.

**Supplementary Materials Figure S2:**
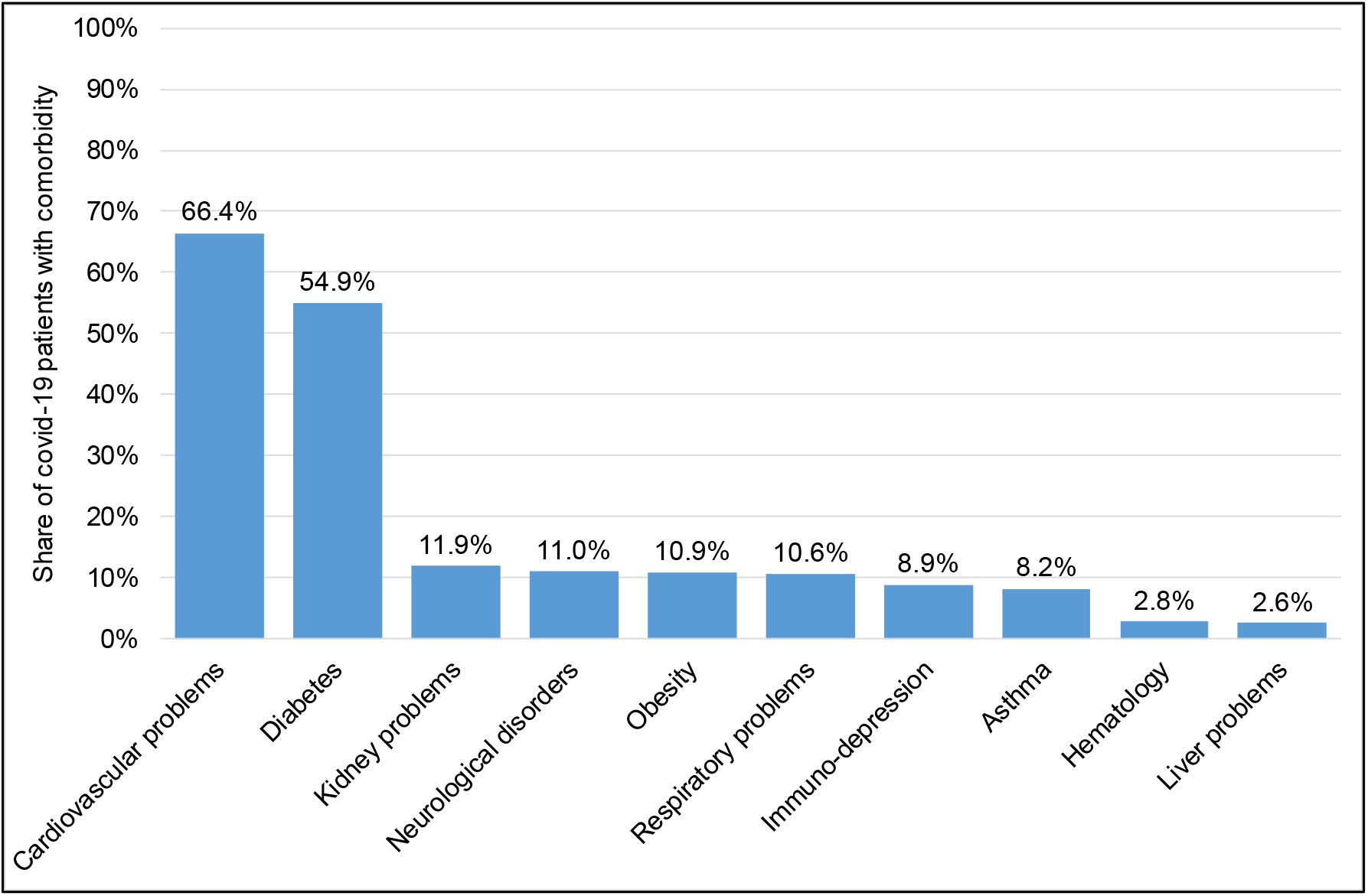
Prevalence of comorbidity among Covid-19 patients. shows the prevalence and type of comorbidities among Covid-19 patients.

**Supplementary Materials Figure S3:**
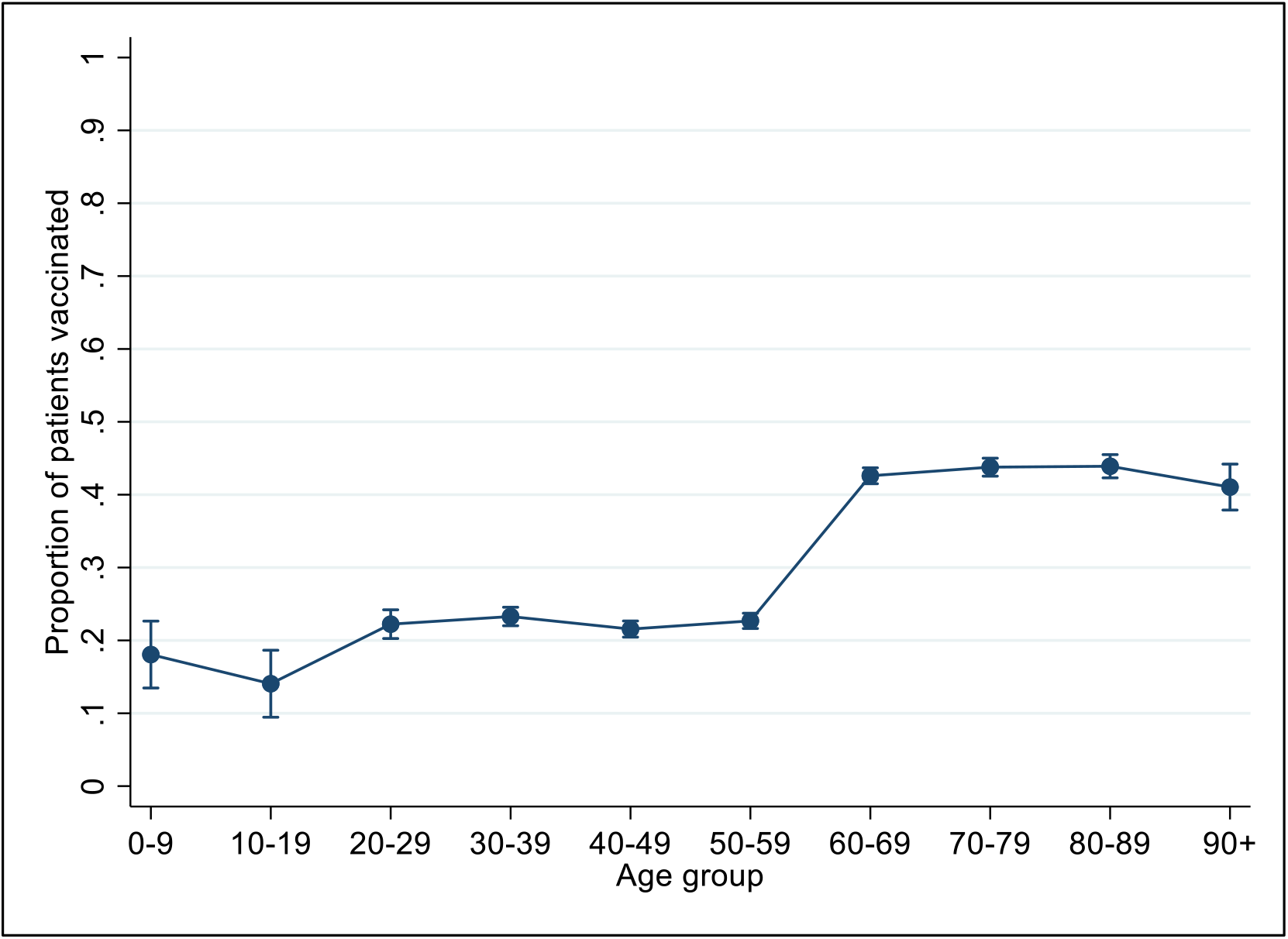
Vaccination coverage by age group. shows age vaccination rates by 10-year age group. Vaccination coverage on the y-axis is expressed as proportion of Covid-19 patients in each age group having received an influenza vaccine in the last 2 years. The 2020 campaign for children started on May 9^th^, reaching only some children by June 8, when the data was extracted.

**Supplementary Materials Figure S4:**
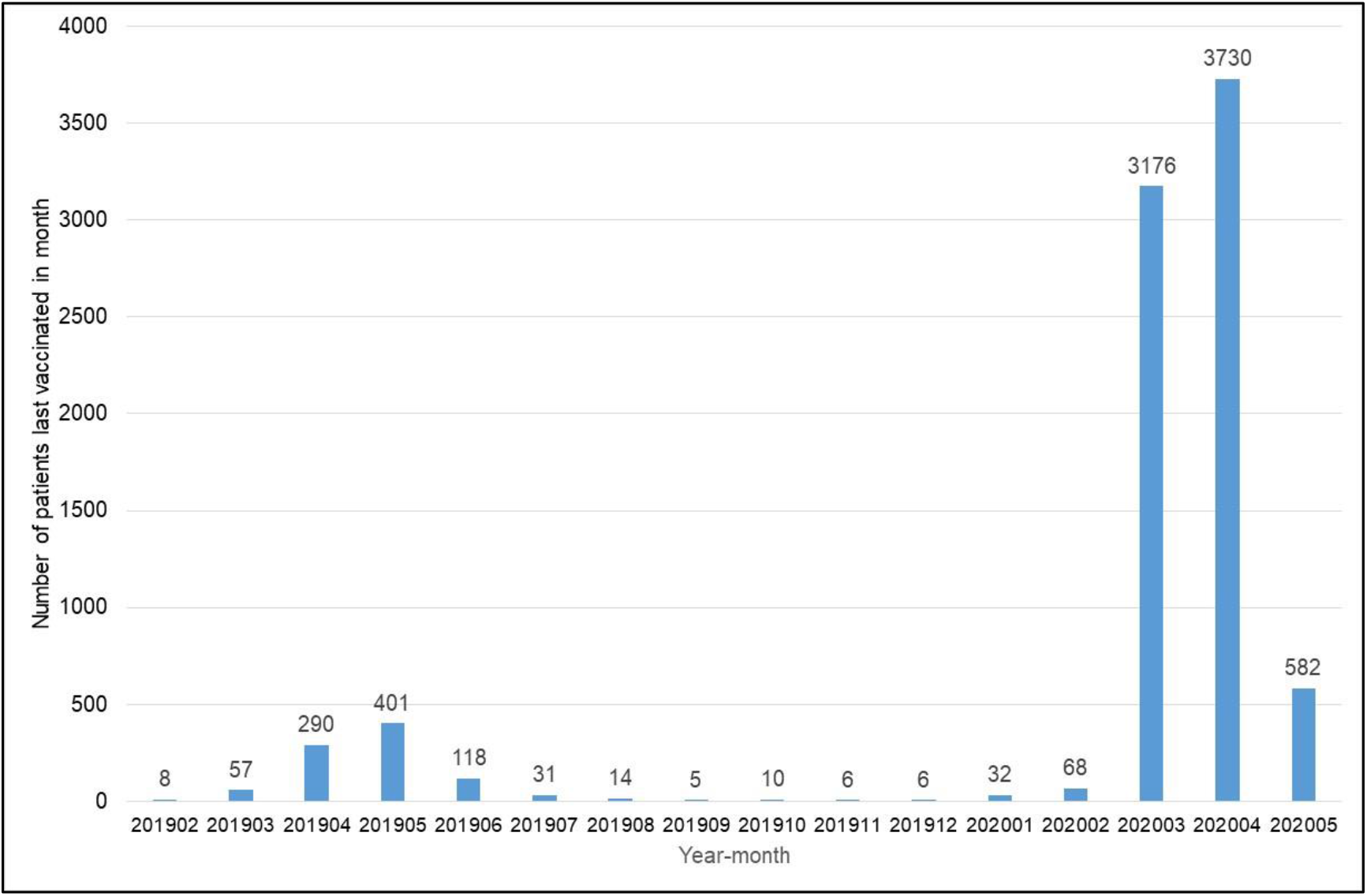
Timing of influenza vaccination. shows the number of (most recently received) influenza vaccines by month of administration.

**Tables S1:**
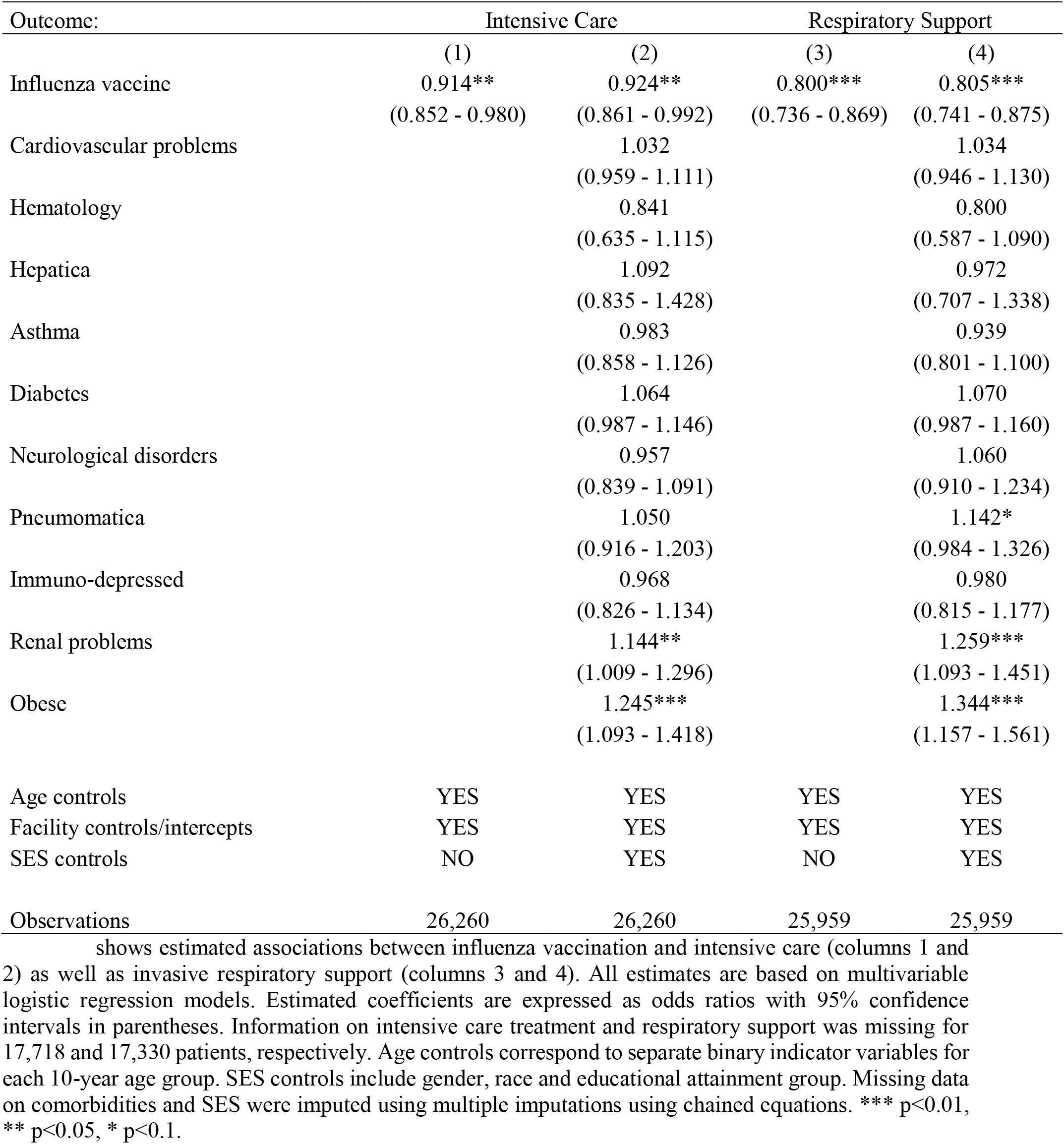
Estimated associations between influenza vaccine and care received (full model)

**Tables S2:**
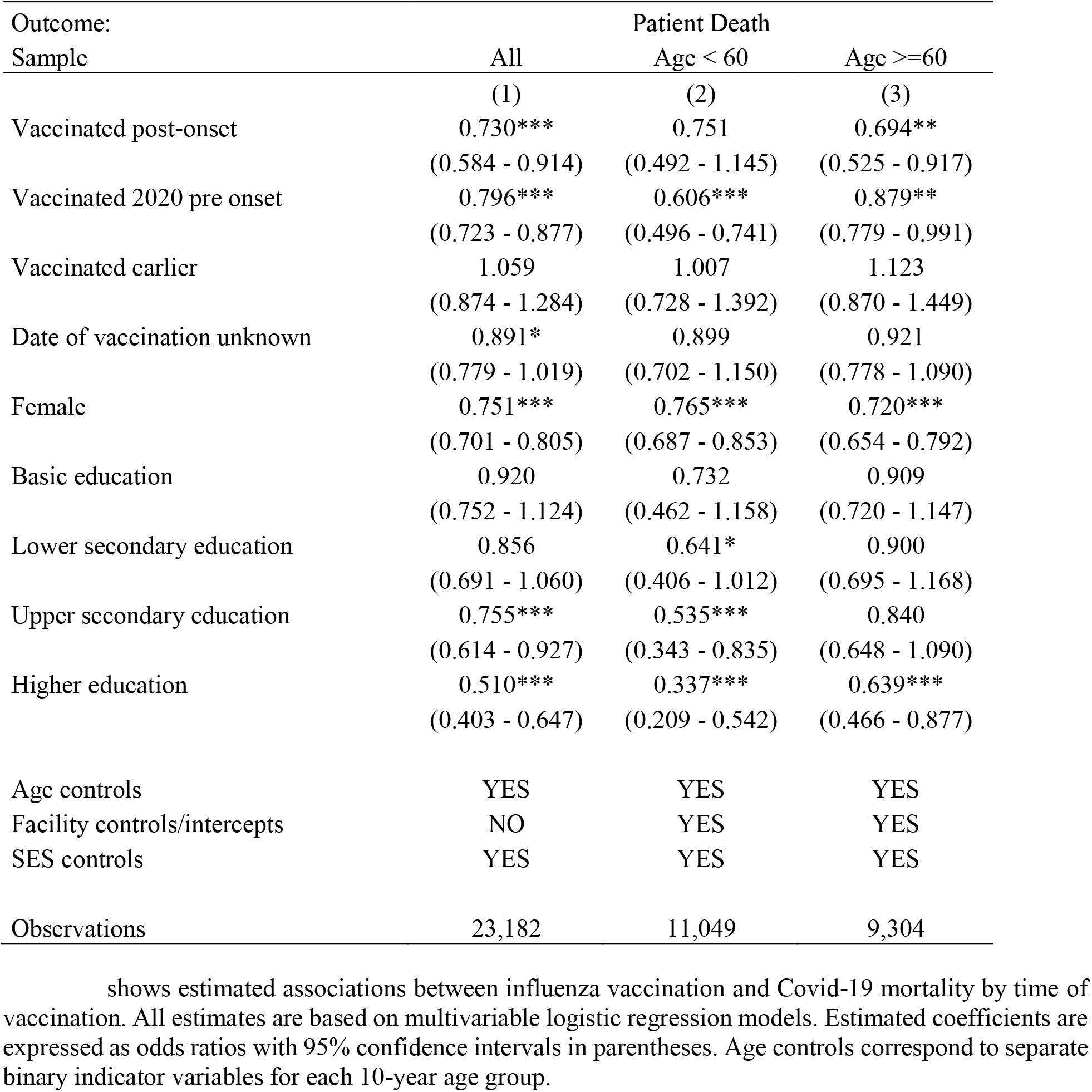
Estimated associations between influenza vaccine and Covid-19 mortality (by time of vaccination)

## References

1. WHO. (WHO, https://www.who.int/emergencies/diseases/novel-coronavirus-2019, 2020), vol. 2020.

2. G. Grasselli et al., Baseline Characteristics and Outcomes of 1591 Patients Infected With SARS-CoV-2 Admitted to ICUs of the Lombardy Region, Italy. Jama 323, 1574–1581 (2020).

3. P. Goyal et al., Clinical Characteristics of Covid-19 in New York City. The New England journal of medicine 382, 2372–2374 (2020).

4. World Bank, Global Economic Prospects. 2020.

5. D. A. Berlin, R. M. Gulick, F. J. Martinez, Severe Covid-19. New England Journal of Medicine, (2020).

6. E. Callaway, Coronavirus vaccine trials have delivered their first results - but their promise is still unclear. Nature 581, 363–364 (2020).

7. qWHO. (2020).

8. P. Schmid, D. Rauber, C. Betsch, G. Lidolt, M. L. Denker, Barriers of Influenza Vaccination Intention and Behavior - A Systematic Review of Influenza Vaccine Hesitancy, 2005 - 2016. PloS one 12, e0170550 (2017).

9. E. Karafillakis, H. J. Larson, The benefit of the doubt or doubts over benefits? A systematic literature review of perceived risks of vaccines in European populations. Vaccine 35, 4840–4850 (2017).

10. Reuters. (https://www.reuters.com/article/uk-factcheck-flu-shot-contains-covid19/false-claim-the-flu-shot-contains-covid-19-idUSKBN22V29N, 2020).

11. A. R. R. Freitas, M. R. Donalisio, Excess of Mortality in Adults and Elderly and Circulation of Subtypes of Influenza Virus in Southern Brazil. Frontiers in Immunology 8, 1903 (2018).

12. S. S. d. Cunha, L. A. B. Camacho, A. C. Santos, I. J. R. d. s. p. Dourado, Influenza vaccination in Brazil: rationale and caveats. 39, 129–136 (2005).

13. A. Almeida, C. Codeço, P. M. Luz, Seasonal dynamics of influenza in Brazil: the latitude effect. BMC Infect Dis 18, 695–695 (2018).

14. B. Ministro da Saúde. (2020).

15. I. Butantan. (Instituto Butantan, 2020).

16. M. J. Cox, N. Loman, D. Bogaert, J. O’Grady, Co-infections: potentially lethal and unexplored in COVID-19. The Lancet Microbe 1, e11 (2020).

17. D. Kim, J. Quinn, B. Pinsky, N. H. Shah, I. Brown, Rates of Co-infection Between SARS-CoV-2 and Other Respiratory Pathogens. JAMA 323, 2085–2086 (2020).

18. X. Wu et al., Co-infection with SARS-CoV-2 and Influenza A Virus in Patient with Pneumonia, China. Emerging infectious diseases 26, 1324–1326 (2020).

19. E. Cuadrado-Payán et al., SARS-CoV-2 and influenza virus co-infection. Lancet (London, England) 395, e84–e84 (2020).

20. M. Wang et al., Clinical diagnosis of 8274 samples with 2019-novel coronavirus in Wuhan. 2020.2002.2012.20022327 (2020).

21. N. Chen et al., Epidemiological and clinical characteristics of 99 cases of 2019 novel coronavirus pneumonia in Wuhan, China: a descriptive study. The Lancet 395, 507–513 (2020).

22. M. D. Nowak, E. M. Sordillo, M. R. Gitman, A. E. Paniz Mondolfi, Co-infection in SARS-CoV-2 infected Patients: Where Are Influenza Virus and Rhinovirus/Enterovirus? n/a.

23. S. A. Plotkin, Updates on immunologic correlates of vaccine-induced protection. Vaccine 38, 2250–2257 (2020).

24. C. H. Lee et al., CD8+ T cell cross-reactivity against SARS-CoV-2 conferred by other coronavirus strains and influenza virus. 2020.2005.2020.107292 (2020).

25. M. G. Netea et al., Defining trained immunity and its role in health and disease. Nature reviews. Immunology 20, 375–388 (2020).

26. M. G. Netea, J. Quintin, J. W. van der Meer, Trained immunity: a memory for innate host defense. Cell Host Microbe 9, 355–361 (2011).

27. M. G. Netea et al., Trained immunity: A program of innate immune memory in health and disease. Science 352, aaf1098 (2016).

28. P. Aaby, T. R. Kollmann, C. S. Benn, Nonspecific effects of neonatal and infant vaccination: public-health, immunological and conceptual challenges. Nat Immunol 15, 895–899 (2014).

29. J. Leentjens et al., BCG Vaccination Enhances the Immunogenicity of Subsequent Influenza Vaccination in Healthy Volunteers: A Randomized, Placebo-Controlled Pilot Study. J Infect Dis 212, 1930–1938 (2015).

30. S. Mohanty et al., Prolonged proinflammatory cytokine production in monocytes modulated by interleukin 10 after influenza vaccination in older adults. J Infect Dis 211, 1174–1184 (2015).

31. L. C. J. de Bree et al., Non-specific effects of vaccines: Current evidence and potential implications. Seminars in immunology 39, 35–43 (2018).

32. J. P. Higgins et al., Association of BCG, DTP, and measles containing vaccines with childhood mortality: systematic review. BMJ 355, i5170 (2016).

33. F. Geeraedts et al., Superior immunogenicity of inactivated whole virus H5N1 influenza vaccine is primarily controlled by Toll-like receptor signalling. PLoS pathogens 4, e1000138 (2008).

34. S. S. Diebold, T. Kaisho, H. Hemmi, S. Akira, C. Reis e Sousa, Innate antiviral responses by means of TLR7-mediated recognition of single-stranded RNA. Science (New York, N.Y.) 303, 1529–1531 (2004).

35. S. Stegemann-Koniszewski et al., Respiratory Influenza A Virus Infection Triggers Local and Systemic Natural Killer Cell Activation via Toll-Like Receptor 7. Front Immunol 9, 245 (2018).

36. H. R. Wagstaffe et al., Influenza Vaccination Primes Human Myeloid Cell Cytokine Secretion and NK Cell Function. J Immunol 203, 1609–1618 (2019).

37. M. K. Andrew et al., Influenza Vaccination in Older Adults: Recent Innovations and Practical Applications. Drugs & aging 36, 29–37 (2019).

38. S. N. Crooke, I. G. Ovsyannikova, G. A. Poland, R. B. Kennedy, Immunosenescence and human vaccine immune responses. Immunity & ageing : I & A 16, 25 (2019).

39. N. Vabret et al., Immunology of COVID-19: Current State of the Science. Immunity, (2020).

40. K. Chumakov, C. S. Benn, P. Aaby, S. Kottilil, R. Gallo, Can existing live vaccines prevent COVID-19? Science 368, 1187–1188 (2020).

41. M. T. Osterholm, N. S. Kelley, A. Sommer, E. A. Belongia, Efficacy and effectiveness of influenza vaccines: a systematic review and meta-analysis. The Lancet. Infectious diseases 12, 36–44 (2012).

